# Physicians’ attitudes and perceived barriers toward ward-based clinical pharmacist in hospital settings: Responses from a national survey

**DOI:** 10.1101/2023.08.08.23293822

**Authors:** Najmaddin A. H. Hatem, Mohammed A. Kubas, Seena A. Yousuf, Abdunaser Rasam, Mohamed Izham Mohamed Ibrahim

## Abstract

Clinical pharmacy services CPSs are still in their infancy in Yemen. Furthermore, pharmacists are not members of a multidisciplinary healthcare team, and their responsibilities are limited to drug dispensing and marketing. Thus, this study investigates physicians’ attitudes and perceived obstacles toward clinical pharmacists working in hospitals’ medical wards. A descriptive observational study was carried out using a validated self-administrated bilingual questionnaire. The study’s questionnaire was conducted among physicians in three teaching hospitals. Those hospitals were at the front to establish clinical pharmacy units and embrace clinical pharmacy services. Sixty-five responses were included. our data results indicated that physicians believed the most contributions for clinical pharmacists to improve patient care was “Attend medical rounds” 70.8%, followed by “Order review”. About 75% of physicians showed positive attitudes toward the clinical pharmacist role. However, more than 70% of physicians thought that clinical pharmacists should leave patient care to other healthcare members and care about drug products. There were Nine potential barriers out of 18 barriers were identified. Not enough clinical pharmacist staff working in the health center was considered as the top perceived barrier 83.1%; followed by clinical pharmacist responsibilities were not clearly defined”, and “clinical pharmacist recommendations are not properly documented”. In terms of enhancing physicians’ general attitudes and overcoming reported barriers. Strategies to expand clinical pharmacy services in Yemen should be emphasized on both, protocols should be established to outline how clinical pharmacists and physicians should collaborate besides, inter-professional collaboration relations are needed to be developed to overcome resistance and raise knowledge and awareness of CPS adoption among the healthcare team members.

## Introduction

Clinical pharmacy is described by the American College of Clinical Pharmacy as the “area of pharmacy concerned with the science and practice of rational drug use” [1]. Clinical pharmacists offer patient-centered services that have evolved to ensure the most effective use of drugs for standard drug therapy [2]. Specifically, those services are designed to optimize therapeutic effect [3,4], minimize risks [5], save cost [6, 7] and put the patient at the forefront of all acts. Clinical pharmacy services CPSs are provided by pharmacists in the healthcare environment in collaboration with physicians and can include counseling and patient education, comprehensive medication management, chronic care management, and population health management for several disease states [8,9]. Clinical pharmacist-physician collaboration in hospital [10] and primary care settings [11,12] have led to improved clinical outcomes for patient with a variety of diseases including osteoporosis [12], diabetes mellitus [13,14,11], hypertension [15,16], hyperlipidemia[17] and infectious disease like HIV and COVID-19 [18,19].

Pharmacists have been demanded to participate in clinical duty and accountability during the last 25 years, specifically with the establishment and application of Hepler and Strand’s [20] concept of the ‘pharmaceutical care’ paradigm. Many developed countries are either in the process of recognizing pharmacists as independent clinical practitioners or have already done so. In the United Kingdom, pharmacists can now not only distribute but also prescribe medications [21]. In Canada, pharmacists assist physicians to design a patient’s prescription regimen [22]. In Australia, pharmacists give consultation services to individuals who have been referred by their general practitioner (GP) [23]. Furthermore, in some healthcare settings in the USA, they are part of a multi-disciplinary intensive care unit team that contributes to the care of patients [24]. However, considering the certain improvements in the pharmacy profession in a few advanced nations, there remains persistent opposition to the expansion of clinical pharmacy practice in both developed and developing countries [25,26,27]. CPSs, for example, have not been extensively utilized in Swedish hospitals. According to studies from Iran, Nigeria, Pakistan and Yemen ineffective clinical training, the view of pharmacists as dispensers by other healthcare practitioners, and a lack of regulation are all factors that negate pharmacists’ clinical tasks [28,29,30,31].

Yemen is an economically low-income developing country in the medial east region. Clinical pharmacy services are still in their infancy in Yemen. Further, pharmacists are not members of a multidisciplinary healthcare team, and their responsibilities are limited to drug dispensing and marketing. Although the current model of direct patient treatment still needs to be completely implemented [32]. However, pharmacists’ functions are evolving to keep up with the demands of a modern healthcare system. whereas, Clinical pharmacy administration was established by the Ministry of Health (MOH) in 2019. Concurrently, the Ministry of Health released the first national clinical pharmacy guideline, outlining job responsibilities and allowing clinical pharmacists to register their interventions as part of clinical pharmacy services. Moreover, 8 colleges are offering clinical pharmacy programs five of them established 5 years bachelor of clinical pharmacy, and the other three colleges provided 6 year Pharm.D program. however, the Ministry of Higher Education and Ministry of Health consider all those who graduated from both programs to be bachelor’s degree holders. In 2011, Hodeidah University became the first university to offer a clinical pharmacy program, and in 2013, it established the first independent college of clinical pharmacy.

Previous research have indicated that there are some variations in how doctors see various types of CPSs [33–43] and that doctors seem to have an improved perception of CPSs if they interact with pharmacists more often [33,37–41]. Doctors in Sweden are sceptical of the expertise and understanding of clinical pharmacists [33]. Moreover, medical students from a Midwestern medical school in the United States do not believe that the task of a pharmacist duties include patient screening and physical examination [44]. Concerns about a pharmacist’s clinical competency continue to exist in Sierra Leone [37]. When it came to advanced clinical pharmacy tasks like interfering in prescriptions and pharmaceutical therapy, consultations, prescribing, etc., people in Pakistan were skeptical and the concept of pharmacists joining an allied healthcare team was not accepted well with the doctors [38].

In conclusion, it is critical to highlight that successful clinical pharmacy service delivery primarily depends on physician collaboration. Hence, exploring physicians’ attitudes toward clinical pharmacists is critical in implementing and moving clinical pharmacy forward. Furthermore, clinical pharmacists face numerous challenges when implementing their services; thus, it is critical to recognize the factors that impede clinical pharmacist interventions, as well as to address those challenges in the future and extend the scope of clinical pharmacist interventions [20]. At the time of writing this article, just three hospitals had introduced clinical pharmacy services and included clinical pharmacists in separate wards. Therefore, such a research has yet to be undertaken to investigate physicians’ attitudes and perceived obstacles to physicians working with ward-based clinical pharmacists in Yemeni hospitals. Further, we aimed to examine whether the attitudes of physicians affected by socio-demographic factors of the respondents.

## Methodology

### Study setting

This study was conducted in three teaching hospitals. Those hospitals were at the front to establish clinical pharmacy units and embrace clinical pharmacy services. At the time of preparing this paper, no other hospitals are providing clinical pharmacy services or employing clinical pharmacists in Yemen. The University of Science and Technology Hospital (USTH) is one of the most prestigious medical institutions in the country. The USTH is located in the capital of Yemen. No clinical pharmacists graduated from Yemeni universities when the clinical pharmacy unit was founded in 2013. Hence, the USTH and 48 Hospital hired clinical pharmacists who earned their master’s degrees in a clinical pharmacy outside of Yemen. The 48 Hospital also located in Sana’a and it is the university hospital of 21 of September university. Althowrah Hospital-Al-Hudaydah (AHA) is the first governmental hospital that started to provide clinical pharmacy services in 2017. (AHA) located in Al-Hudaydah city. Which is nearly 217 Km far from the capital. It began by hiring only those with a Bachelor’s degree in clinical pharmacy from Hodeidah University. Since it was the first governmental university to launch a clinical pharmacy program, it has been a leader in the field.

### Study Design and study tool

A descriptive observational study was carried out from September to November 2021 using a self-administrated questionnaire developed to determine the physician attitudes and barriers toward clinical pharmacists in two teaching hospitals. The questionnaire was adopted from previous quantitative studies with minor modifications. The questionnaire will be introduced in both Arabic and English language. The questionnaire included both closed and open-ended questions. It consists of three parts. The socio-demographic information of physicians’ Age, gender, Years of experience in practice, and current area of practice were represented in the first part. In the second part, the 5-Likert scale of (Strongly agree = 5, Agree = 4, Neutral = 3, Disagree = 2, and Strongly disagree = 1) was used to assess physicians’ attitudes by using 16 questions adopted from previous studies [41–43]. The third part was concerned with the barriers that may hinder the clinical pharmacists’ contributions to the health care team those questions were adopted from existent literature [34,35]. Two independent academic members from the faculty of pharmacy pre-tested the questionnaire for suitability and content relevance. The questionnaire was additionally reviewed by other three physicians for relevance, clarity, conciseness, and simplicity of the items. The questionnaire’s final version included feedback from experts.

### Ethical approval

All participants who took part in the study gave their verbal permission. The Deanship of College of Clinical Pharmacy at Hodeidah University, Al-Hudaydah, Yemen accepted the study’s design and methodology in compliance with the Declaration of Helsinki with Reference No. (DM-2-1255).

### Data Collection

After receiving permission from the hospital’s administrators. Questionnaires were distributed by three well-trained and non-pharmacy-related majors. They handled the questionnaires to physicians. And due to the physician’s nature of work the questionnaires were left with them, and then after three days responses were gathered and forwarded to the lead researchers. The study was conducted between July 2021 to February 2022.

### Data Analysis

To analyze the data, they were first entered and coded in Excel, then imported into SPSS version 26.0 (SPSS, Chicago, IL, USA). To summarize categorical data, basic descriptive statistics including frequency, percentage, mean, and standard deviations were used. Cronbach’s alpha was used to measure the overall reliability and internal consistency of the questionnaire. Its total value was 0.831, and each item scored more than 0.8. During the data analysis attitude’s items were categorized to (positive/negative) were attitude’s item that has a Mean of 4 or more was considered positive attitude’s item. While, in barriers items were categorized to (Potential/non-potential barriers), were barrier that has a Mean of 3 or more considered potential barrier. To examine the relationship between a dependent variable (attitude and barriers) and demographic factors (age, gender, occupation, and years of experience), logistic regression was used at an alpha level of 0.05.

## Results

Sixty-five responses were included (60% response rate). In Table 1. more than three-quarters of them 76.9% were males, and more than half were under 40 years old. Nearly 37% of physicians were specialists, and the majority have experience more than 15 years of practice. About 31% work in the internal medicine ward. Around two-thirds of the participants either had “Never” 12.3% or “Rarely” 21.5% had working experience with a clinical pharmacist. 40% of physicians rat that clinical pharmacist services provided in the included hospitals were “Acceptable” in terms of meeting the patient’s needs. Also, our data results indicated that physicians believed the most contributions for clinical pharmacists to improve patient care were “Attend medical rounds” 70.8%, followed by “Order review” and “Patient education” 58.5%, and 52.3% respectively more information can be found in Table 2.

**Table 1.** Socio-demographic information of participant physicians.

| Variables | N | % |
| --- | --- | --- |
| **Sex** |  |  |
| Male | 50 | 76.9 |
| Female | 15 | 23.1 |
| **Age (years)** |  |  |
| <30 | 15 | 23.1 |
| 30-39 | 20 | 30.8 |
| 40-49 | 21 | 32.3 |
| ≥50 | 9 | 13.8 |
| **Professional role** |  |  |
| Consultant | 12 | 18.5 |
| Specialist | 24 | 36.9 |
| Resident | 12 | 18.5 |
| GP | 17 | 26.2 |
| **Experience (years)** |  |  |
| 0-3 | 18 | 27.7 |
| 4-7 | 11 | 16.9 |
| 8-11 | 5 | 7.7 |
| 12-15 | 7 | 10.8 |
| >15 | 24 | 36.9 |
| **Area of Practice** |  |  |
| Internal Medicine | 20 | 30.8 |
| Obstetrics and gynecology | 7 | 10.8 |
| Critical Care | 3 | 4.6 |
| Pediatrics | 8 | 12.3 |
| Surgery | 17 | 26.2 |
| Other | 10 | 15.4 |
| **Type of Hospital** |  |  |
| Governmental | 25 | 38.5 |
| Private | 21 | 32.3 |
| Both | 19 | 29.2 |

**Table 2.** Physicians’ working experience, perception, and contribution area of ward-based clinical pharmacist.

| Variables | N | % |
| --- | --- | --- |
| **Experience working with clinical Pharmacist** |  |  |
| Never | 8 | 12.3 |
| Rarely | 14 | 21.5 |
| Sometimes | 30 | 46.2 |
| Often | 10 | 15.4 |
| Always | 3 | 4.6 |
| **Clinical pharmacist meets the needs in terms of patient care** |  |  |
| Not at all | 9 | 13.8 |
| Poor | 10 | 15.4 |
| Acceptably | 26 | 40.0 |
| Adequately | 16 | 24.6 |
| Very adequately | 4 | 6.2 |
| **Most important contribution for clinical pharmacist to improve patient care** |  |  |
| Attend medical rounds | 46 | 70.8 |
| Being available for consultation | 32 | 49.2 |
| Patient education | 34 | 52.3 |
| Order review | 38 | 58.5 |
| Staff education | 27 | 41.5 |
| Attend clinic | 15 | 23.1 |

In Table 3. About 75% of physicians showed positive attitudes toward clinical pharmacists with a mean and stander deviation of 4.06±0.43. More than 90% of physicians positively welcomed the clinical pharmacist to participate in the medical ward round, “monitor patients’ responses to drug therapy from a toxicity/side effects perspective” and “believed that clinical pharmacists can improve the overall patient outcome/quality of patient life”. Moreover, 89.2% of physicians positively thought that “clinical pharmacists can provide drug information to other healthcare team members regarding compatibility, stability, storage, and availability”. However, more than 70% of physicians thought that clinical pharmacists should leave the patient care to other healthcare members and care about drug products, and a good number of physicians negatively thought “clinical pharmacist can be involved in drug selection”, and “Clinical pharmacist should analysis patient and treatment and suggest changes of therapy when necessary”. While the statement “the current setup appropriate for the provisions of clinical pharmacy services” received the lowest agreement 49.3%; around 29% of physicians were “Neutral”.

**Table 3.** Attitudes of physicians toward ward-based clinical Pharmacist role.

| Statement | Strongly agree | Agree | Neutral | Disagree | Strongly disagree | Mean ± SD | Attitude (positive/negative) |
| --- | --- | --- | --- | --- | --- | --- | --- |
| Clinical pharmacist participation in medical ward round is desirable. | 31 (47.7) | 28 (43.1) | 4 (6.2) | 2 (3.1) | -- | 4.35 ± 0.73 | Positive attitude |
| Clinical pharmacist can play important role in patient education and counseling. | 28 (43.1) | 26 (40.0) | 11 (16.9) | -- | -- | 4.26 ± 0.73 | Positive attitude |
| Clinical pharmacist can monitor patient response to drug therapy from toxicity/side effects perspective. | 29 (44.6) | 31 (47.7) | 4 (6.2) | 1 (1.5) | -- | 4.35 ± 0.67 | Positive attitude |
| Clinical pharmacist can monitor patient response to drug therapy from effectiveness perspective. | 18 (27.7) | 33 (50.8) | 12 (18.5) | 2 (3.1) | -- | 4.03 ± 0.77 | Positive attitude |
| Clinical pharmacist can involve in drug selection (drug, dosage form) based on patient and drug factors. | 14 (21.5) | 31 (47.7) | 15 (23.1) | 2 (3.1) | 3 (4.6) | 3.78 ± 0.97 | Negative attitude |
| Clinical pharmacist can provide drug information to health care professionals such as compatibility, stability, storage, availability. | 22 (33.8) | 36 (55.4) | 7 (10.8) | -- | -- | 4.23 ± .63 | Positive attitude |
| Clinical pharmacist can detect and prevent medication use errors. | 20 (30.8) | 37 (56.9) | 7 (10.8) | 1 (1.5) | -- | 4.17 ± 0.67 | Positive attitude |
| Clinical pharmacy service enhances patients' appreciation and satisfaction. | 18 (27.7) | 33 (50.8) | 12 (18.5) | 2 (3.1) | -- | 4.03 ± 0.77 | Positive attitude |
| Clinical pharmacist should take patients medication history at admission. | 25 (38.5) | 30 (46.2) | 8 (12.3) | 2 (3.1) | -- | 4.20 ± 0.77 | Positive attitude |
| Clinical pharmacist should have access to patients chart and have a place to document their service. | 26 (40.0) | 26 (40.0) | 10 (15.4) | 1 (1.5) | 2 (3.1) | 4.12 ± 0.94 | Positive attitude |
| Clinical pharmacist should analyses patient treatment and suggest changes of therapy when necessary. | 20 (30.8) | 30 (46.2) | 9 (13.8) | 3 (4.6) | 3 (4.6) | 3.94 ± 1.03 | Negative attitude |
| Clinical pharmacist should care about drug products and leave patient care to Doctors, Health Officers and nurses. | 16 (24.6) | 31 (47.7) | 9 (13.8) | 7 (10.8) | 2 (3.1) | 3.80 ± 1.03 | Negative attitude |
| The current setup (infrastructure and environments of your hospitals) appropriate for the provisions of clinical pharmacy service. | 2 (3.1) | 30 (46.2) | 19 (29.2) | 9 (13.8) | 5 (7.7) | 3.23 ± 0.99 | Negative attitude |
| Clinical pharmacy service initiation is desirable in Yemen health care system. | 20 (30.8) | 33 (50.8) | 10 (15.4) | -- | 2 (3.1) | 4.06 ± 0.86 | Positive attitude |
| Clinical pharmacist can improve overall patient outcome /quality of patient care. | 24 (36.9) | 36 (55.4) | 4 (6.2) | 1 (1.5) | -- | 4.28 ± 0.65 | Positive attitude |
| Overall attitude (mean ± SD) | 4.06 ± 0.43 | | | | | | |

Among the 18 barriers that were included in the study’s questionnaire (Table 4.), there were nine potential barriers were identified. Not enough clinical pharmacist staff working in the health center was considered as the top perceived barrier 83.1%; followed by clinical pharmacist responsibilities were not clearly defined”, “clinical pharmacist recommendations are not properly documented”, and “Administration does not supportively enough clinical pharmacy services” 81.5%, 53.8%, and 52.3% respectively. While the statement that clinical pharmacist needs more confidence to interact with other healthcare team received the lowest agreement to be a potential barrier 32.5%.

**Table 4.** Barriers that can hinder ward-based clinical pharmacist’s role with physicians.

| Statement | Strongly agree | Agree | Neutral | Disagree | Strongly disagree | Mean ± SD | Potential/ non-potential barrier |
| --- | --- | --- | --- | --- | --- | --- | --- |
| The specific responsibilities of a clinical pharmacist are not clearly defined. | 16 (24.6) | 37 (56.9) | 10 (15.4) | 2 (3.1) | -- | 4.03± 0.73 | Potential barrier |
| There are not enough number of clinical pharmacists staff in the health center that we are working in. | 16 (24.6) | 38 (58.5) | 5 (7.7) | 6 (9.2) | -- | 3.98± 0.84 | Potential barrier |
| Administration does not supportively enough clinical pharmacist services. | 12 (18.5) | 22 (33.8) | 22 (33.8) | 7 (10.8) | 2 (3.1) | 3.54± 1.02 | Potential barrier |
| The hospital that we work in is inconvenient. | 4 (6.2) | 12 (18.5) | 17 (26.2) | 25 (38.5) | 7 (10.8) | 2.71± 1.08 | Non-potential barrier |
| Clinical pharmacist does not have good knowledge on disease management. | 4 (6.2) | 19 (29.2) | 23 (35.4) | 11 (16.9) | 8 (12.3) | 3.00± 1.10 | Potential barrier |
| Clinical pharmacist does not have good knowledge on drug related issues. | 4 (6.2) | 11 (16.9) | 22 (33.8) | 19 (29.2) | 9 (13.8) | 2.72± 1.10 | Non-potential barrier |
| Clinical pharmacist does not have enough confidence to interact with the health care team. | 6 (9.2) | 21 (32.3) | 15 (23.1) | 18 (27.7) | 5 (7.7) | 3.38± 2.60 | Potential barrier |
| Clinical pharmacist is not interested to provide clinical pharmacy services. | 3 (4.6) | 16 (24.6) | 15 (23.1) | 22 (33.8) | 9 (13.8) | 2.72± 1.12 | Non-potential barrier |
| Clinical pharmacists have inadequate communication skills needed for interaction with physicians. | 3 (4.6) | 20 (30.8) | 18 (27.7) | 16 (24.6) | 8 (12.3) | 2.91± 1.11 | Non-potential barrier |
| Clinical pharmacist recommendations are not properly documented. | 8 (12.3) | 27 (41.5) | 17 (26.2) | 12 (18.5) | 1 (1.5) | 3.45± 0.98 | Potential barrier |
| There is no continuity in the services provided by the clinical pharmacist due to other non-clinical roles. | 2 (3.1) | 26 (40.0) | 24 (36.9) | 11 (16.9) | 2 (3.1) | 3.22± 0.92 | Potential barrier |
| Physicians are unaware of the benefits of having a clinical pharmacist on their team. | 4 (6.2) | 26 (40.0) | 10 (15.4) | 19 (29.2) | 6 (9.2) | 3.05± 1.15 | Potential barrier |
| Physicians are unable to judge the knowledge and level of skills of the clinical pharmacist. | 6 (9.2) | 20 (30.8) | 12 (18.5) | 19 (29.2) | 8 (12.3) | 2.95± 1.21 | Non-potential barrier |
| Physicians have low trust in the clinical pharmacist's abilities. | 1 (1.5) | 16 (24.6) | 19 (29.2) | 19 (29.2) | 10 (15.4) | 2.68± 1.06 | Non-potential |
|  |  |  |  |  |  |  | barrier |
| Physicians have inadequate communication skills needed for interaction with the clinical pharmacist. | -- | 13<br>(20.0) | 19<br>(29.2) | 21 (32.3) | 12<br>(18.5) | 2.51±<br>1.01 | Non-potential barrier |
| Physicians do not support adequately the clinical pharmacist services. | -- | 15<br>(23.1) | 22<br>(33.8) | 17 (26.2) | 11<br>(16.9) | 2.63±<br>1.02 | Non-potential barrier |
| Physicians have no prior experience of working with a clinical pharmacist. | 7 (10.8) | 23<br>(35.4) | 13<br>(20.0) | 17 (26.2) | 5 (7.7) | 3.15±<br>1.16 | Potential barrier |
| Physicians believe that clinical pharmacists cannot be clinicians. | 5 (7.7) | 14<br>(21.5) | 13<br>(20.0) | 21 (32.3) | 12<br>(18.5) | 2.68±<br>1.22 | Non-potential barrier |
| Overall barriers (mean ± SD) | 3.07 ± 0.66 |  |  |  |  |  |  |

## Discussion

This is the first observational study to highlight how clinical pharmacist roles are perceived by physicians with regards to their contribution to direct patient care and their participation in the healthcare team furthermore, identify important barriers that could hinder the role of clinical pharmacist in healthcare settings from a physician’s point of view. The vast majority of physicians positively welcomed the clinical pharmacist’s participation in the medical ward round and monitoring patients’ response. However, more than 70% of physicians thought that clinical pharmacists should leave patient care to other healthcare members and care about traditional tasks. The current study revealed nine potential barriers were identified. Not enough clinical pharmacist staff working in the health center and “clinical pharmacist responsibilities were not clearly defined”, were considered as the top two perceived barriers reported by physicians working in the three hospitals.

Clinical pharmacy services CPSs do exist in the sampled hospitals. However, clinical pharmacy is an entirely novel concept in the country’s existing healthcare environment, and CPSs in those hospitals are still in their early stages, confined to medication related problem-solving and providing clinical suggestions for physicians in certain medical wards [3]. Physicians who responded to our survey rated clinical pharmacist services provided in the included hospitals as “Acceptable” in terms of meeting the patient’s needs. Also, our data results indicated that physicians believed that pharmacists can improve patient’s life by performing extended patient-oriented roles such as attending medical rounds, order reviews, patient education, and being available for consultation. These findings are consistent with prior studies [36–38].

### Attitudes aspect

The results demonstrate that physicians show positive attitudes toward clinical pharmacists with a mean and stander deviation of 4.06 ± 0.43. The vast majority of physicians positively welcomed “the clinical pharmacist’s participation in the medical ward round, monitor patients” response to drug therapy from a toxicity/side effects perspective, and believe that clinical pharmacists can improve the overall patient outcome/quality of patient life”. Moreover, they positively thought that clinical pharmacists can provide drug information to other healthcare team members. In the current study, physicians in Yemen encourage the clinical pharmacist’s involvement in the clinical ward team, who may routinely seek guidance on patient medication. This conclusion is consistent with the findings of several earlier investigations [36,37,39]. In addition, physicians revealed that clinical pharmacists may enhance overall health outcomes and patient quality of life. These views are in agreement with those expressed earlier by Saudis and US doctors, who pointed out that clinical pharmacists’ participation and recommendations increased the quality of patient care [40,45]. A recent studies found that clinical pharmacist intervention and engagement in clinical decisions can reduce hospital stays, medication errors, patient satisfaction, and the impact of quality of care [46,47,48,49].

However, the results from our study demonstrate that physicians view traditional-tasks such as providing drug information regarding compatibility, stability, storage, and availability to be the responsibility of clinical pharmacists. While, the vast majority of them reported clinical pharmacists should leave the patient care to other healthcare members and care about drug products. This was expected since many low-income nations reported the responsibilities of the clinical pharmacist have been defined as subordinate to that of the physicians [35,50,51]. One probable explanation for these perceptions is that physicians are unaware of clinical pharmacists’ expertise. When the clinical pharmacist and the physician continue to communicate and the pharmacist can share his/her expertise, physicians may see clinical pharmacists’ potential and significance to the health care team. International experience suggests that physicians’ attitudes and acceptance of pharmacists’ clinical duties have improved over time and have been attributed to physicians’ interaction with pharmacists and continuous exposure to their services, with these negative attitudes often fading away [52,53,54,35]. Consequently, to maintain a successful working environment, a clear line must be placed between these skills and those of other medical professionals. In this country, the recently developing pharmacy curriculum included some instructions on analyzing patients’ signs and symptoms, including physical examination and complete assessments of patients’ biomarker profiles to determine patients’ medication demands [32,55]. These capacities are shared by other medical practitioners. Pharmacists as clinicians are not widespread in Yemen pharmacy practice [32]. Hence, some physicians are in doubt of their skills and prefer their conventional dispensing orientation.

Additional explanation is lack of inter-professional education (IPE), which is driven by the fact that there is lately no established protocol outlining how clinical pharmacists and physicians should collaborate. Hence, inter-professional collaboration relations are needed to be developed to overcome resistance and raise knowledge and awareness of CPS adoption among the healthcare team members. Therefore, defining and describing each healthcare professional’s tasks and responsibilities is crucial while creating a multidisciplinary healthcare team. Additionally, during both doctors’ and pharmacists’ undergraduate training programs, a collaborative or multidisciplinary work environment needs to be supported or fostered.

This study underlines the need of clinical pharmacists interacting effectively and often with physicians on the ward level in order to improve physicians’ impressions of the role of CPSs in Yemen hospitals. It has also been revealed that decision makers’ support for CPSs enhanced implementation. [56]. Hence, some schemes for incentives should be established to encourage clinical pharmacists for rational therapeutic decision-making and to deeply discuss medication selections during ward rounds with physicians.

Numerous research investigations have reported that with the changing level of pharmacy practice in recent years, younger physicians and physicians who have more exposure to pharmacists’ services in the hospital setting appeared to have higher expectations and attitudes toward pharmacists and were more accepting of pharmacists’ new patient-focused roles [38,56]. In the current, we did not find any significant difference that supports this idea or any significant difference among other physicians’ Socio-demographic information that was included in this study. Evidence was also in line with previous studies conducted [37,57,58].

### Barriers aspects

Among the eighteen barriers that were included in the study’s questionnaire, there were nine potential barriers were identified. When trying to carry out clinical tasks, pharmacists face challenges such as a lack of time, a shortage of staff, and a heavy workload [56,57,36]. According to the report released in 2019, the total number of pharmacists in Yemen reached ≈ 18,000 having one pharmacist for every 1700 [58]. Additionally, pharmacy technicians’ professional actions are not regulated by any national or institutional policies. The evidence suggests that pharmacy technicians can assist pharmacists with logistical tasks, giving them time to spend on other patient care-related issues and to engage in more clinical activities [55].

The current investigation provided evidence that the duties of clinical pharmacists are not well defined. Many studies conducted in other countries shared the same reported barrier [56,58,36]. Similar to this, in 2009 team members in a Canadian study of healthcare organizations pointed out that the lack of a defined role for pharmacists results in misconceptions and barriers that limit the provision of CPSs [49]. Currently, no clear model care has included clinical pharmacists in the healthcare system in Yemen and the traditional model care is the predominant model which is a triangle-like shape clearly stating that physicians are the only in clinical decision-making.

Therefore, implementing clinical pharmacy services necessitates the integration of a new cross-disciplinary collaboration model where pharmacists engage in clinical decision-making, as well as the introduction of a new profession into providing of healthcare. Additionally, different parts of the world where CPSs are being implemented for the first time in the nation’s health system, new care models and the measures that correlate with them should be used [6,38,50]. As a result, increasing contact is required to increase awareness of CPSs and their advantages. When the task of pharmacist clearly established, his performance in multidisciplinary patient-care teams improved. According to a previous investigation, an official statement of the duties assigned to each team member assists in promoting awareness of individual commitments and minimize conflict that arises from a lack of established limits [37,38,40]. Hence, the growing acceptability of clinical pharmacists and their pharmaceutical care interventions, which improve the standard of healthcare, could continue if physicians’ and nurses’ awareness of clinical pharmacy services and attitudes toward the profession significantly improved [37,38,39,40,59].

Clinical pharmacists can take on ever-expanding duties including managing patients’ pharmacotherapy and therapeutic drug monitoring, both of which considerably enhance patient care [20,61]. The nature of their contacts affect how well clinical pharmacists and doctors work together to provide care to patients [13,60]. Clinical pharmacists may answer questions on a range of topics, such as drug profiles, dosages, adverse drug reactions, patient management, drug interactions, drug usage during pregnancy and lactation, poisons, and suggestions for drug storage [6,7,9]. They collaborate with patients and other healthcare providers to encourage and help patients adjust their lifestyles to achieve better health results. Improvements in patient care, level of awareness, and illness management have the effect of reducing risk factors and healthcare costs [10,12,13].

### Limitations

The relatively low response rate of 65 assessed surveys, which was lower than our projected response rate, is a drawback of our study. This was most likely affected by the research population’s heavier workload. Another concern is the social desirability bias: respondents may have given favorable comments to adhere to the more socially accepted viewpoint. Furthermore, because the survey was conducted at a given time and certain hospitals, it does not account for any long-term changes in physicians’ attitudes, or perceived barriers concerning ward-based clinical pharmacists and their services. Despite these drawbacks, we think these findings offer insightful information about how doctors feel about clinical pharmacists and the barriers preventing them from offering their recently introduced services.

## Study implication on practice and recommendations

When the clinical pharmacist and the physician continue to communicate and the pharmacist can share his/her expertise, physicians may see clinical pharmacists’ potential and significance to the health care team. Consequently, to maintain a successful working environment, a clear line must be placed between these skills and those of other medical professionals. International experience suggests that physicians’ attitudes and acceptance of pharmacists’ clinical duties have improved over time and have been attributed to physicians’ interaction with pharmacists and continuous exposure to their services, with these negative attitudes often fading away [52,53,54,35]. Further, ward-based pharmacists are still uncommon in Yemen, as previously said, and many doctors and nurses are not aware of the competencies that clinical pharmacists can preformed. The largest number of pharmacists continue to work in community pharmacies, where giving patients advice and collaborating with other health professionals are still their less preformed responsibilities in Yemen [46]. Hence, implementing clinical pharmacy services necessitates the integration of a new cross-disciplinary collaboration model where pharmacists engage in clinical decision-making, as well as the introduction of a new profession into providing of healthcare, in other words a protocol should be established to outline how clinical pharmacists and physicians should collaborate and inter-professional collaboration relations are needed to be developed to overcome resistance and raise knowledge and awareness of CPS adoption among the healthcare team members. Additionally, during both doctors’ and pharmacists’ undergraduate training programs, a collaborative or multidisciplinary work environment needs to be supported or fostered.

## Conclusions

In Yemen, physicians appreciate the role of clinical pharmacists in direct patient care but they preferred that clinical pharmacists focus more on drug product services such as drug information sources and patient education, while limiting drug selection and treatment change for physician responsibilities. A limited number of clinical pharmacists staff in hospitals and unclear definitions of clinical pharmacist roles were the most common barriers reported by Yemeni physicians. Protocols should be established to outline how clinical pharmacists and physicians should collaborate and inter-professional collaboration relations are needed to be developed to overcome resistance and raise knowledge and awareness of CPS adoption among the healthcare team members. Additionally, during both doctors’ and pharmacists’ undergraduate training programs, a collaborative or multidisciplinary work environment needs to be supported or fostered.

## Data Availability

All relevant data are within the manuscript and its Supporting Information files.

## Acknowledgment

The second author would like to thank USTH for their financial support which allow him to copy and distribute the questionaries’ in the hospital.

